# Stigma, Place, and HIV: The Legacy of the 1984 Bhopal Gas Disaster Among Indian Men Who Have Sex with Men

**DOI:** 10.64898/2026.10.31.26366861

**Authors:** Griffin J Bell, Carly A Comins, Jason M Lo Hog Tian, Allison M McFall, Eshan U. Patel, Talia A Loeb, Mihili P Gunaratne, Jiban J Baishya, Ashwini Kedar, Archit Sinha, AK Srikrishnan, Boobalan Jayaseelan, Gregory M. Lucas, Shruti H Mehta, Sunil S Solomon, Steven J Clipman

**Author notes:** **Corresponding Author:** Griffin J Bell.

## Abstract

**Background:** The 1984 Bhopal Gas Leak is widely considered the worst industrial disaster in history, and “big events” can have durable impacts on infectious disease transmission. Among men who have sex with men, we examined whether meeting sexual partners near the original gas leak site was associated with HIV, HIV-associated risk behaviors, and sexual stigma.

**Methods:** We used respondent-driven sampling to enroll MSM in Bhopal during 2023-24 (n=750). Participants reported the four locations where they most often went to meet sexual partners in the past year. Sexual stigma was defined by mechanism (enacted, anticipated, internalized) and source (healthcare, family, society). We calculated weighted (RDS-II) prevalences of HIV, population-level HIV viremia (≥200 copies/ml), and associated risk behaviors by meeting location type and distance to the original gas leak site. We used weighted logistic regression to examine associations between sexual stigma and partner meeting locations.

**Results:** HIV prevalence was elevated among participants meeting near the leak site: 21.7% within 1 km, 14.4% between 1-2 km, and 9.7% over 2 km. Population-level HIV viremia and the proportion of MSM engaging in sex work were also elevated within 2 km. Sexual stigma measures were significantly associated with meeting a partner within 2 km of the leak site. Transportation-related spaces were associated with higher prevalences of HIV and viremia.

**Conclusions:** Industrial disasters may create marginal spaces which concentrate individuals facing greater stigma and elevated HIV transmission risk. Venue-based interventions near the gas leak site and at transportation centers may be warranted in Bhopal.

**Funding:** This study was supported by the National Institute on Drug Abuse grant R01DA041034. GJB, CC, and TL were supported by National Institute of Allergy and Infectious Diseases grant T32AI102623. SJC was funded by the National Institute on Drug Abuse grant DP2DA056130. SSS was funded by National Institute on Drug Abuse grant DP1DA060602.

## Introduction

Large-scale events and policies are increasingly recognized as structural drivers of HIV transmission. Systematic economic disinvestment in Black communities in the United States and South Africa, facilitated through policies of residential segregation, created environments of concentrated poverty and limited healthcare infrastructure, which still impact the spatial distribution of HIV infection and viral suppression outcomes decades after those policies ended (1–3). For people who inject drugs, “big events” such as economic recessions and the COVID-19 pandemic are recognized as key drivers of HIV outbreaks, shaping injection networks and risk behaviors (4–6). Similarly, armed conflicts, like in Ukraine, can accelerate HIV transmission through population displacement and mixing of injection and sexual networks, reshaping the geospatial distribution of infections (7). These events and policies often operate through place-based mechanisms, reshaping physical and social structures in ways that concentrate harm among already-marginalized populations and embed effects that persist for decades.

The literature on the legacy effects of industrial and natural disasters on HIV and other infectious diseases is limited. One study found elevated incidence rates of tuberculosis in areas more affected by the Chernobyl disaster, and noted that the results were probably not a direct impact of radiation but rather due to a “set of negative factors that have a pernicious influence on the general health of the population” (8). Studies have linked hurricanes and other extreme weather events to short-term disruptions in HIV services and increases in HIV risk behaviors, especially among vulnerable populations (9–14), though few studies have captured how these events influenced the burden of HIV over the following decades.

The Bhopal Gas Disaster is widely regarded as the worst industrial disaster in human history (15,16). On December 2^nd^, 1984, approximately 40 tons of methyl isocyanate gas leaked from the Union Carbide pesticide plant in Bhopal, India, engulfing the surrounding neighborhoods, including informal settlements. Approximately 10,000 people died within three days of the leak, and more than 25,000 deaths have since been attributed to the disaster (15,16). In total, over half a million people were exposed to the gas (16). Long-term health consequences, including greater risk of chronic respiratory disease, ocular issues, immunotoxicity, neurological and psychiatric conditions, and adverse reproductive and birth outcomes, have been documented even amongst subsequent generations (15,17–20). Significant contamination around the plant persists, with groundwater showing elevated levels of heavy metals and chlorinated organic pollutants (21), but the last comprehensive contamination report was completed over a decade ago (22). The response to the disaster was widely criticized as inadequate. Union Carbide’s 1989 settlement of 470 million USD, which amounted to 25,000 rupees (approximately 500 USD) per victim, was insufficient, and ongoing efforts to seek justice from the individuals responsible have proven unsuccessful (16). To date, there has been no comprehensive operation to remove the toxic waste from the site (23). Like many large-scale events and policies, the impact of the Bhopal disaster has been spatially and socially heterogeneous, with disproportionate burden falling upon impoverished communities close to the original leak site (16,20). While many long-term consequences of the Bhopal disaster have been extensively documented, its potential role in shaping the burden of HIV has not been examined.

Overall HIV incidence in India has been steadily declining (24,25), yet HIV burden among key populations, including men who have sex with men (MSM), remains disproportionately high (26): an estimated 2.4% of MSM were living with HIV compared to 0.2% among all adults aged 15-49 years (27). A study conducted across 10 Indian cities from 2016-17 using RDS found a weighted HIV prevalence of 17.6% among MSM overall and 16.3% among MSM in Bhopal (28). Indian MSM face unique sociocultural and psychosocial pressures and remain a largely hidden population (29). Same-sex relations between men were criminalized under Section 377 of the Indian Penal Code until 2018, and its legacy continues to shape the social and structural context, including access to education and income-generating activities and vulnerability to poverty and violence (30). Approximately 30.6% of MSM in India are married to a woman (31), with many reporting societal stigma and familial pressures to marry, have children, and conceal their sexual practices. These social pressures contribute to experiences of living a double life, elevated rates of alcohol dependence (32), adverse childhood experiences, depression, and suicidality (33). MSM in India further contend with discrimination, internalized stigma, and enacted stigma by family members and healthcare providers (33,34), influencing healthcare access, utilization, and retention in care.

Stigma and structural exclusion also influence sexual behaviors among MSM, including the locations where MSM find and meet partners (35). Among MSM in India, compounding pressures of stigma and social marginalization channel partner-seeking into hidden, semi-public spaces with documented venues including parks, public toilets, bus and railway stations, among others (36). Risk tolerance, or the willingness to accept situations or behaviors that may increase exposure to HIV/STI infection, violence, or other negative experiences, is often influenced by personal beliefs and life context (37–40). Thus, within stigmatizing settings, desire to remain hidden and risk tolerance to engaging in same-sex practices may increase. Venue typology influences HIV risk and vulnerability, shaping condom negotiation power, access to outreach services, and exposure to violence (41). A subset of MSM in India engage in transactional sex, defined as the buying or selling of sex or sexual services for money or goods, and experience compounded HIV risk and increased stigma, discrimination, and structural marginalization (42,43). The physical, social, and geographic locations where MSM meet other men could inform venue-based HIV prevention and treatment interventions, potentially among underserved MSM unreachable through clinic-based interventions.

Large-scale events and policies have been shown to act through characteristics of place to durably concentrate HIV epidemics within specific vulnerable or stigmatized communities. Thus, the enduring impacts of the Bhopal disaster may influence the sexual partner meeting locations and distribution of HIV among MSM in Bhopal. Here, we map sexual partner meeting locations among MSM in Bhopal to 1) identify the characteristics of meeting locations frequented by all MSM and MSM with HIV to inform HIV control programs and venue-based interventions, 2) explore the relationship between proximity to the original 1984 gas leak site and HIV prevalence and viremia, and 3) identify any differences in sexual behaviors and sexual stigma among MSM meeting partners near the leak site.

## Methods

### Study Design

Enrollment for this cross-sectional study of MSM in Bhopal, India, was done between 2023 and 2024 using respondent-driven sampling (44,45), which is commonly used for sampling key populations where no sampling frame exists (46). Recruitment began with the selection of 2 seeds who were well connected and influential among MSM in Bhopal. Seeds were given two paper recruitment coupons to distribute to other MSM whom they knew and asked to encourage recipients to attend the study site and present the coupon. Those eligible were enrolled and given two recruitment coupons. This iterative peer recruitment continued until the desired sample size of 750 (excluding seeds) was reached. Eligibility criteria included: at least 18 years old, self-identified as male, reported oral or anal sex with a man in the prior 12 months, and presented a valid RDS referral coupon (except seeds). People who identified as women, transgender, or hijra were excluded. All enrolled participants provided a blood sample and completed an interviewer-administered electronic survey including sections on socio-demographics, sexual behavior, uptake of HIV treatment and prevention interventions, and stigma. Biometrics (i.e., iris) were used to ensure people did not enroll twice in the study. Participants were compensated for completing RDS study procedures (300 Indian rupees (INR)) and for each eligible and enrolled participant they referred (100 INR), up to two total.

### HIV Status and Viremia

HIV status was determined through on-site rapid testing and counseling under Indian standards. Participants were provided with pre- and post-test counseling, and those living with HIV were referred to HIV care/treatment at local government HIV care/treatment centers. Plasma HIV-1 RNA viral loads were determined using the Abbott RealTime HIV-1 assay (Abbott Park, IL, USA), and participants were considered virally suppressed with viral loads <200 copies/mL and viremic otherwise. Population prevalence of viremia was computed using all participants regardless of HIV status as the denominator.

### Survey Data

In the survey, participants reported up to four areas of Bhopal where they had most often gone to meet sexual partners in the past 12 months by providing a place name and approximate location using Google Maps on a tablet. Participants also reported socio-demographic and sexual behavior characteristics. Sociodemographic variables included age (years), sexual orientation (heterosexual, homosexual, bisexual), marital status (never married, currently married, widowed or divorced), education (primary, secondary or high school, post-secondary), wage earning pattern (monthly or weekly, daily, intermittent employment, not employed, student), average monthly personal income in the last 12 months (rupees), living situation (own, rent home, rent room, other), number of children, running out of basics (e.g. food) in the last 12 months (yes, no), sleeping outside in the last 12 months (yes, no), and access to a smartphone (yes, no). Sexual partner variables corresponded to the previous six months and included number of regular partners, number of casual partners, and condom use among regular and casual partners. Here, we examined use only among casual partners (consistent/perfect, inconsistent, never, no casual partners), as condom use among casual partners was expected to be a more relevant determinant of transmission risk.

Guided by established stigma frameworks (47–49), we measured multiple dimensions of sexual stigma using a structured 25-item survey capturing three stigma mechanisms (enacted, anticipated and internalized) across various sources (healthcare workers, family, and society). Items were adapted for the Indian context, informed by existing stigma scales (49–52), and supported by existing qualitative data indicating that healthcare workers, family, and society are salient sources of stigma for MSM and other key populations in India (33,53,54). The survey included seven subscales: *enacted* healthcare stigma (3 items; α=0.70), *enacted* family stigma (3 items; α=0.88), *enacted* societal stigma (4 items; α=0.87), *anticipated* healthcare stigma (3 items; α=0.71), *anticipated* family stigma (3 items; α=0.76), *anticipated* societal stigma (4 items; α=0.86), and *internalized* stigma (5 items; α=0.71). *Enacted* stigma items paralleled *anticipated* stigma items, where *enacted* stigma items assessed the frequency of past discriminatory experiences and *anticipated* stigma assessed perceived likelihood of future mistreatment from a given source for being MSM. *Internalized* stigma measured current negative feelings about being MSM. Responses were recorded on Likert scales (enacted stigma: 0 = never to 3 = frequently; anticipated stigma: 0 = very unlikely to 3 = very likely; internalized stigma: 0 = not at all to 3 = a great deal), and mean scores were calculated for each subscale (theoretical range: 0-3), with higher scores indicating greater stigma. Individual survey questions can be found in the Supplemental Methods.

### Spatial Data Processing

To identify unique sexual partner meeting locations reported by MSM, all reported locations were first cleaned and deduplicated before final geocoding. First, we identified groups of named destinations likely representing the same location and assigned a unique and consistent place name for all observations representing that location. We then geocoded those unique names with the Google Maps API. We compared those geocoded locations to those reported by participants attending that location. When the location obtained through the Google Maps API differed substantially from participant-reported locations, these errors were resolved manually. Most locations (303/327; 92.7%) were accurately geocoded by Google API after name cleaning and harmonization, while the remaining 24 locations were geocoded by hand. We categorized each location into one of commercial (shop, market, mall, cineplex), neighborhood (hotel, house, unknown spot in neighborhood), transportation-related (bus stop/terminal, train stop/station, road, fuel pump), park (park, fairgrounds, trail, wedding grounds), and other (e.g., school, temple, factory, hospital).

### Statistical Analyses

“Seed” participants were removed from all analyses. RDS-II weights were created using the inverse of reported network size (55). Network sizes were determined by the answer to the question “Of the [total number of MSM known by the participant] people who live in Bhopal, who you personally know are MSM, how many have you seen at least once in the past 30 days?” We calculated weighted summary statistics and used the tree bootstrap method (100,000 replicates) to calculate 95% confidence intervals (56). We summarized characteristics overall and stratified by HIV status using both RDS-II weighted and unweighted percentages for categorical variables, and medians and first and third quartiles for continuous variables. We also calculated the prevalence of experiencing any sexual stigma (i.e., score>0) across mechanisms and sources, stratified by HIV status. We calculated the Euclidean distance from each meeting place to the original gas leak site and categorized participants according to their closest reported meeting place into the following categories: less than 1 kilometer (km), between 1 and 2 km (inclusive), and over 2 km. We then calculated summary statistics stratified by distance, as well as the weighted prevalence of HIV and HIV viremia. Because reporting more partner meeting locations may be associated with increased sexual activities (and therefore HIV), while also providing additional opportunities to meet a partner near the 1984 site, we conducted a sensitivity analysis where we restricted to participants who reported the maximum of four meeting locations to recalculate the HIV prevalence gradient across distance to the leak site.

Finally, we fit univariable weighted logistic regression models to estimate the odds of meeting a sexual partner within 2 km (dichotomized for modeling simplicity) of the gas leak site. We used sexual stigma measures as the exposure, and a separate regression was fit for each stigma type and source. We conducted a sensitivity analysis where we repeated our logistic regression analyses, but restricting only to people who did not live within 2 km of the leak site, to ensure that our results are not just identifying correlations among those living and meeting within that radius, but also among people traveling to the area. For both main and sensitivity analyses, we also fit adjusted logistic regressions to account for potential confounding by age, sexual orientation, marital status, education, personal income (rupees per month divided by 10,000), and HIV status.

### Ethics

This study was approved by the Johns Hopkins Medicine (IRB 00082575) and the YR Gaitonde Centre for AIDS Research and Education (IRB 00001423) Institutional Review Boards; all participants provided oral informed consent.

## Results

### Participant Characteristics

Of 750 MSM enrolled in the study, the overall weighted HIV prevalence was 11.6% (95% CI: 6.9%-17.9%), and the weighted population prevalence of HIV viremia was 4.4% (95% CI: 1.4%-8.5%). The median age was 24 years (IQR 21-30), with participants living with HIV tending to be older (median 30; IQR 24-38) (Table 1). Almost every participant lived in Bhopal for over a year (99.8%) and completed secondary school (99.5%). Though income did not vary widely by HIV status, participants living with HIV were more likely to identify as homosexual (66.9% vs 49.8%), own a home (79.4% vs. 68.8%), and to have slept outside (16.6% vs 7.9%) in the last 12 months. However, only one participant reported being currently homeless. Overall, 97.8% had at least one casual sexual partner in the last six months, and 40.7% reported ten or more casual partners. Reporting ten or more casual partners was more common among participants with HIV (64.4% vs 37.6%). Condom use with casual partners was typically inconsistent (87.6%). Over two-thirds reported selling sex or exchanging sex for goods (67.1%), and 29.6% bought or exchanged goods for sex. Almost half (45.1%) of those living without HIV had been tested for HIV, but only 8.4% had tested in the last year. Unweighted data are available in Supplemental Table 1.

**Table 1:** Characteristics of men who have sex with men in Bhopal recruited using respondent-driven sampling in 2023-2024, by HIV status (RDS-II weighted)

| Characteristic<br>Median/interquartile range; n/% | **Overall<br>(N=750)** | **With HIV<br>(n=116)** | **Without HIV<br>(n=634)** |
|---|---|---|---|
| **Socio-Demographics** |  |  |  |
| Age | 24 (21, 30) | 30 (24, 38) | 23 (21, 29) |
| Sexual Orientation |  |  |  |
| *Heterosexual* | 216 (43.0) | 22 (28.7) | 194 (44.9) |
| *Homosexual* | 488 (51.8) | 87 (66.9) | 401 (49.8) |
| *Bisexual* | 46 (5.2) | 7 (4.4) | 39 (5.3) |
| Lived in Bhopal >=1 year | 749 (99.8) | 116 (100.0) | 633 (99.8) |
| Marital Status |  |  |  |
| *Never Married* | 531 (73.5) | 64 (56.2) | 467 (75.8) |
| *Currently Married* | 214 (26.1) | 50 (42.4) | 164 (23.9) |
| *Widowed or Divorced* | 5 (0.4) | 2 (1.4) | 3 (0.3) |
| Highest Education |  |  |  |
| *Primary* | 8 (0.5) | 4 (3.0) | 4 (0.2) |
| *Secondary or High School* | 204 (21.7) | 49 (34.5) | 155 (20.0) |
| *Post-Secondary* | 538 (77.8) | 63 (62.5) | 475 (79.7) |
| Earning Pattern |  |  |  |
| *Monthly or Weekly Wages* | 348 (41.2) | 53 (41.8) | 295 (41.1) |
| *Daily Wages* | 84 (8.4) | 30 (23.9) | 54 (6.4) |
| *Intermittently Employed* | 3 (0.2) | 2 (1.3) | 1 (0.1) |
| *Not Employed* | 23 (2.1) | 7 (5.4) | 16 (1.7) |
| *Student* | 292 (48.1) | 24 (27.7) | 268 (50.8) |
| Personal income (Indian rupees) | 40000 (35000, 50000) | 40000 (31414, 45000) | 40000 (35000, 50000) |
| Housing Status |  |  |  |
| *Own* | 557 (70.0) | 96 (79.4) | 461 (68.8) |
| *Rent (Home)* | 114 (14.1) | 16 (13.0) | 98 (14.2) |
| *Rent (Room)* | 69 (13.5) | 3 (5.6) | 66 (14.6) |
| *Other (staying with family, in staff quarters, or in a slum)* | 10 (2.4) | 1 (2.0) | 9 (2.4) |
| Number of Children |  |  |  |
| *0* | 557 (76.5) | 76 (65.0) | 481 (78.0) |
| *1* | 60 (8.3) | 11 (8.9) | 49 (8.3) |
| *2-4* | 133 (15.2) | 29 (26.1) | 104 (13.8) |
| Ran out of basic needs (e.g., food) in last 12 months | 126 (12.5) | 24 (16.0) | 102 (12.0) |
| Slept outside at least one night in last 12 months | 127 (8.9) | 29 (16.6) | 98 (7.9) |
| Access to smartphone | 740 (98.5) | 114 (98.6) | 626 (98.5) |
| **Sexual Behaviors** |  |  |  |
| Number of Regular Partners (last 6 months) |  |  |  |
| *0* | 102 (7.0) | 18 (9.6) | 84 (6.7) |
| *1* | 181 (37.2) | 16 (17.1) | 165 (39.8) |
| *2* | 312 (40.9) | 50 (49.8) | 262 (39.7) |
| *3+* | 155 (14.9) | 32 (23.5) | 123 (13.7) |
| Number of Casual Partners (last 6 months) |  |  |  |
| *0* | 8 (2.2) | 1 (2.0) | 7 (2.2) |
| *1* | 44 (13.6) | 1 (0.6) | 43 (15.3) |
| *2-4* | 163 (32.3) | 10 (17.0) | 153 (34.3) |
| *5-9* | 82 (11.1) | 13 (15.9) | 69 (10.5) |
| *10+* | 453 (40.7) | 91 (64.4) | 362 (37.6) |
| Condom Use with Casual Partners |  |  |  |
| *Consistent* | 64 (7.7) | 9 (6.7) | 55 (7.9) |
| *Inconsistent* | 636 (87.6) | 97 (84.2) | 539 (88.0) |
| *Never* | 42 (2.5) | 9 (7.0) | 33 (1.9) |
| *No Casual Partners* | 8 (2.2) | 1 (2.0) | 7 (2.2) |
| Sell Sex | 560 (67.1) | 102 (85.7) | 458 (64.7) |
| Buy Sex | 292 (29.6) | 63 (50.5) | 229 (26.9) |
| **HIV Treatment and Prevention** |  |  |  |
| Ever tested for HIV | 430 (48.9) | 96 (78.0) | 334 (45.1) |
| HIV Test in the Last Year | 98 (8.1) | 10 (5.7) | 88 (8.4) |
| Knowledge of HIV-Positive Status | - | 33 (21.7) | - |
| Currently taking ARVs | - | 32 (21.1) | - |
| Viral Suppression (<200 copies/ml) | - | 78 (62.1) | - |
| **Experiencing Any Sexual Stigma (score>0)^** |  |  |  |
| Enacted Healthcare | 148 (10.5) | 32 (19.7) | 116 (9.3) |
| Enacted Family | 124 (8.2) | 32 (19.4) | 92 (6.7) |
| Enacted Society | 122 (7.9) | 22 (13.0) | 100 (7.2) |
| Anticipated Healthcare | 731 (98.9) | 112 (98.4) | 619 (98.9) |
| Anticipated Family | 734 (98.9) | 114 (97.7) | 620 (99.1) |
| Anticipated Society | 734 (98.8) | 111 (96.0) | 623 (99.1) |
| Internalized | 237 (19.8) | 44 (27.3) | 193 (18.8) |
| **Sexual Stigma score, Mean (SD)** |  |  |  |
| Enacted Healthcare | 0.07 (0.25) | 0.18 (0.42) | 0.06 (0.21) |
| Enacted Family | 0.08 (0.29) | 0.18 (0.43) | 0.07 (0.27) |
| Enacted Society | 0.07 (0.26) | 0.12 (0.35) | 0.06 (0.24) |
| Anticipated Healthcare | 1.03 (0.33) | 1.04 (0.42) | 1.02 (0.31) |
| Anticipated Family | 1.19 (0.50) | 1.29 (0.61) | 1.17 (0.49) |
| Anticipated Society | 1.06 (0.39) | 1.10 (0.53) | 1.06 (0.37) |
| Internalized | 0.11 (0.28) | 0.17 (0.34) | 0.10 (0.27) |

*^More than “never” (enacted),”very unlikely” (anticipated), and “not at all” (internalized) for any subscale item*

### Sexual Partner Meeting Locations

Participants reported 2,715 sexual partner meeting locations, and 94% of participants reported three or four locations (mean 3.6 per participant). This corresponded to 327 unique places, including 189 neighborhoods, 63 transportation-related locations, 29 commercial locations, 28 parks and grounds, and 18 other locations. Most (n=291, 89.0%) locations were outside of the 2-kilometer radius of the original site of the gas leak, but the two most reported transportation-related locations were between 1-2 km of the site (Figure 1). Participants reporting four locations were more likely to report a meeting place within 2 km of the gas site compared to participants reporting three (39.4% vs. 28.4%).

**Figure 1:**
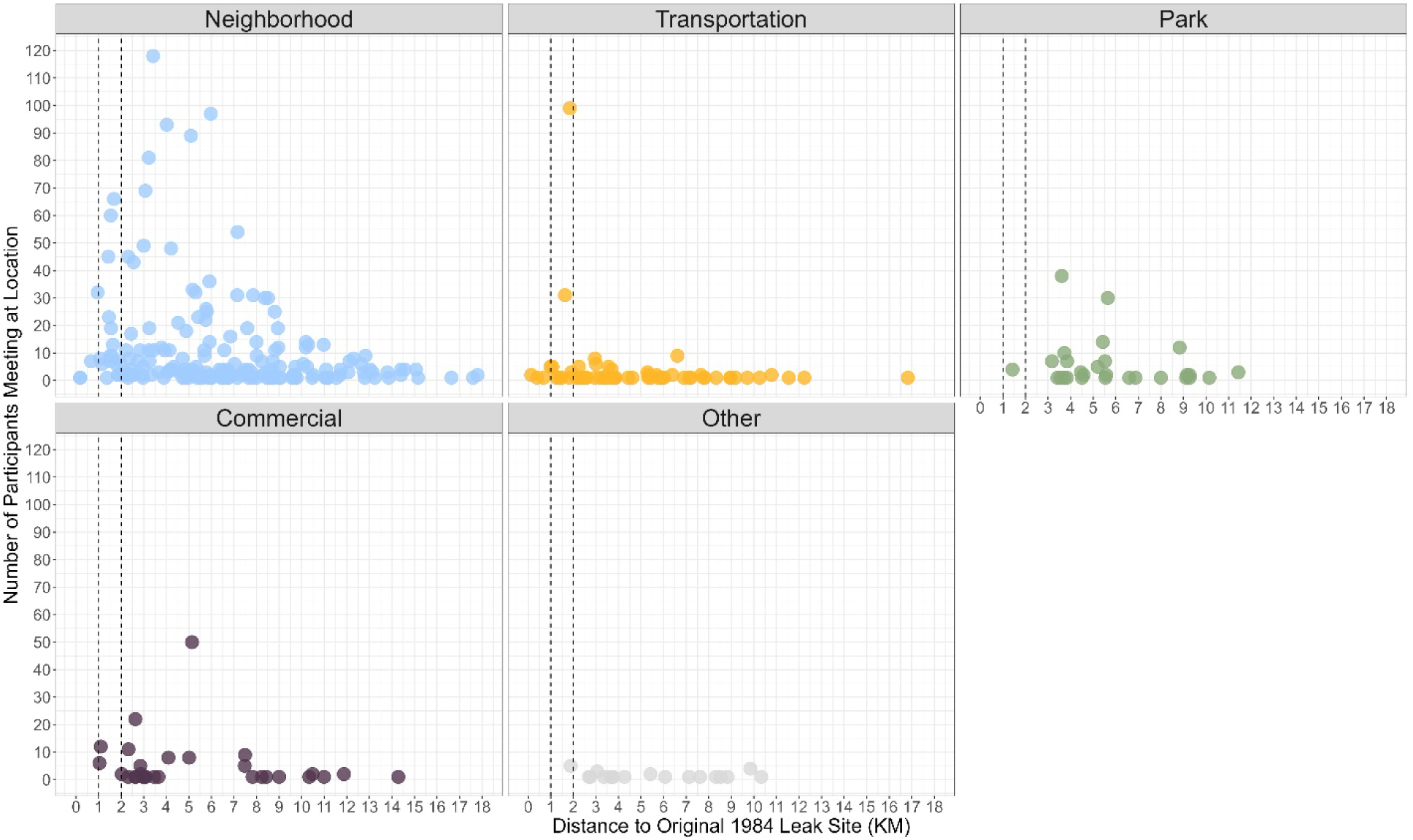
Locations Where Participants Met Men for Sex, Characterized by Distance to the Original 1984 Gas Leak Site and the Number of Participants Reporting the Location. *Shown here are the sexual partner meeting locations reported by participants, characterized by distance to the original 1984 gas leak site and the number of participants that reported it. Each point represents a single location, and are colored by location type (neighborhood, transportation-related, park, commercial, other). A traditional map of these locations is not shown for privacy and ethical reasons. Four neighborhoods (one reported twice, the others reported once) >18 km away were excluded from the plot to increase legibility*.

HIV prevalence was highest among participants meeting partners at transportation-related locations (14.8%, 95% CI: 2.7%-26.4%) and lowest at commercial locations (6.8%, 1.7%-15.7%), compared to 11.4% (6.4%-18.7%) for neighborhoods and 10.4% (2.5%-24.6%) for parks. A similar result was observed for HIV viremia: 6.4% (0.0%-15.8%) for transportation-related vs. 3.5% (0.0%-11.9%) for commercial, 4.3% (1.1%-8.8%) for neighborhoods, and 4.5% (0.0%-16.1%) for parks (Figure 2). HIV prevalence was elevated when a participant met sexual partners near the leak site: 21.7% (2.4%-58.8%) within 1 km, 14.4% (6.1%-24.3%) between 1-2 km, and 9.7% (5.0%-17.1%) over 2 km away (Figure 3). HIV viremia was elevated among participants meeting within 1 km (7.4% [0.0%-32.2%]) and between 1-2 km (6.9% [1.0%-17.1%]) compared to those meeting over 2 km away (3.2% [0.5%-7.3%]). These gradients remained after restricting to only participants reporting the maximum of four meeting locations: 34.3%, 15.9%, and 11.1% for HIV prevalence and 12.1%, 7.3%, 3.6% for HIV viremia for <1, 1-2, and >2 km, respectively. When stratifying by distance to the leak site, HIV prevalence at transportation-related locations was similar to neighborhoods within 2 km of the leak site (17.5% vs. 15.8%) and to neighborhoods and parks beyond 2 km (10.5% vs. 9.4% and 11.0%, respectively) (Figure 2).

**Figure 2:**
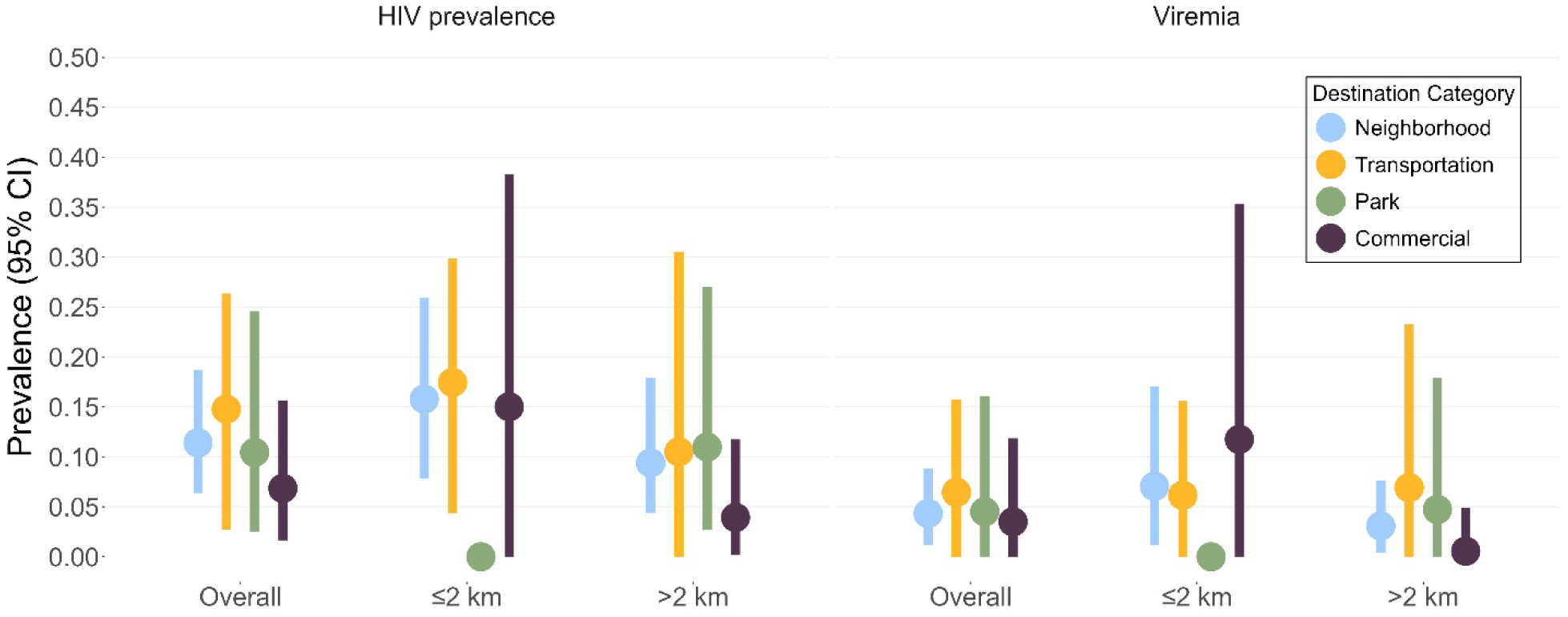
Weighted HIV and Viremia Prevalence and 95% Confidence Interval by Destination Type Overall and Stratified by Distance to the Original Leak Site. *Shown here are HIV prevalence and population prevalence of viremia (proportion of all MSM that are viremic, regardless of HIV status). HIV and viremia prevalences are RDS-II weighted, confidence intervals are calculated using a tree bootstrap (100,000 replicates)*.

**Figure 3:**
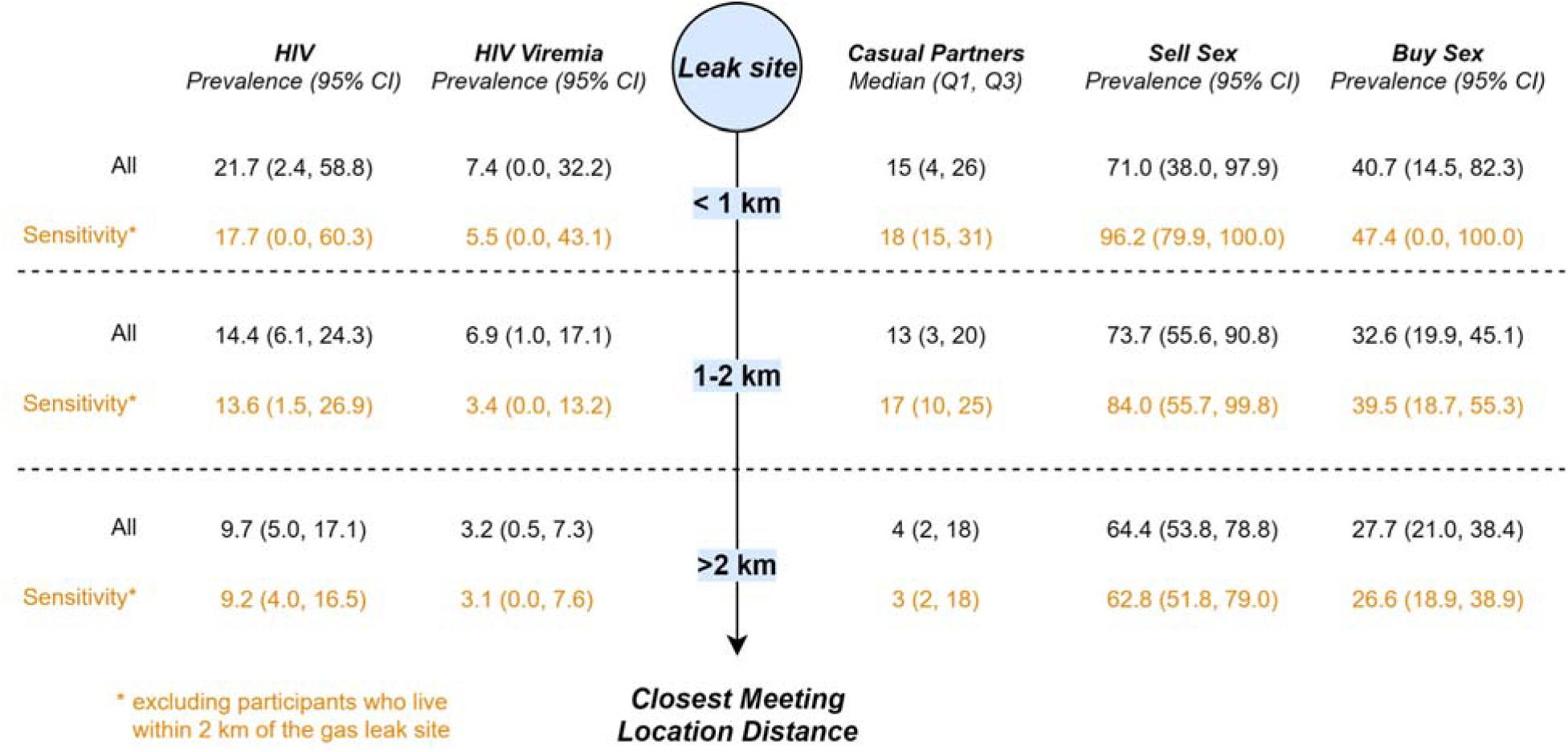
HIV Prevalence, Population Prevalence of Viremia, and Sexual Behaviors by Closest Meeting Site Distance from the Original Gas Leak Site. *Shown here are the RDS-II weighted prevalences of HIV, HIV viremia, selling sex, and buying sex, as well as the 25^th^, 50^th^, and 75^th^ percentiles for casual partner counts, stratified by distance of the participant’s closest partner meeting location to the original gas leak site. Confidence intervals are calculated using a tree bootstrap (100,000 replicates). All participants are included in the main analysis (black) while only participants living outside 2km are included in the sensitivity analysis (orange), to ensure that results are not just due to the characteristics of participants both living and meeting near the leak site*.

In addition to elevated HIV and HIV viremia, 71.0% of participants meeting within a kilometer of the site reported exchanging sex for goods, compared to 73.7% of those meeting within 1-2 km and 64.4% of those only meeting more than 2 km away (Figure 3). Fewer people reported exchanging goods for sex, but the probability also decreased the further the meeting location was from the leak site (Figure 3). Those meeting within 2 km of the leak site also reported more casual partners (57.9%, 57.0%, 33.5% reporting 10+ partners in <1, 1-2, and >2 km distance groups, respectively), as well as elevated proportions of sleeping outside in the last 12 months (15.0%, 19.6%, 4.5%) (Supplemental Table 2). Unweighted comparisons between those meeting within 2 km and outside 2 km are available in the supplement (Supplemental Table 3).

### Sexual Stigma and Sexual Partner Meeting Locations

Anticipated stigma was more common than enacted or internalized stigma, regardless of source, but enacted and internalized stigma showed greater differences by HIV status (Table 1). All stigma measures, regardless of source, were significantly associated with meeting within 2 km of the leak site (Figure 4). Most notably, a one-point increase in internalized stigma was associated with 5.1 (3.2-8.5) times the odds of meeting near the site. A one-point increase in experiencing enacted stigma in a healthcare setting (4.2, 2.6-7.2), from family members (3.8, 2.4-6.1), and from other members of society (3.8, 2.3-6.4) was also strongly associated. Anticipated stigma, whether in a healthcare setting (1.8, 1.2-2.7), from family members (1.6, 1.3-2.1), and from other members of society (1.4, 1.0-1.9), had a weaker, but still significant, association. In our sensitivity analysis restricting to only participants living outside the 2 km radius, the associations remained statistically significant and consistent in direction, though with point estimates further from the null (Odds Ratio = 1) and wider confidence intervals (Figure 4). Adjustment for potential confounders generally resulted in point estimates closer to the null, with a smaller impact on the main analyses compared to sensitivity analyses. Adjusted confidence intervals newly crossed the null for anticipated stigma from family and society sources in both main and sensitivity analyses, but remained statistically significant for all other stigma types and sources (Figure 4).

**Figure 4:**
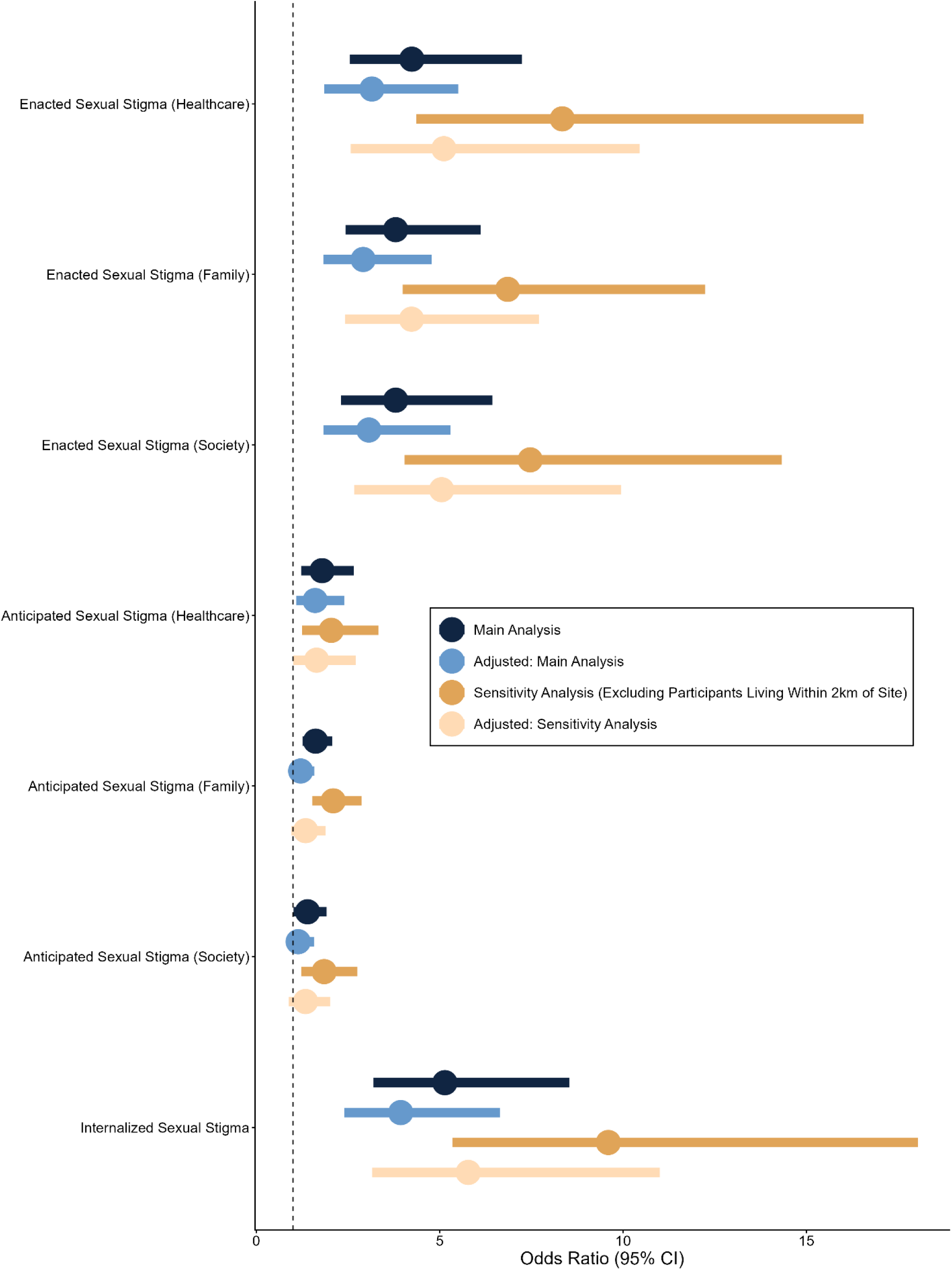
Sexual Stigma and Odds of Meeting ≤2km from Leak Site. *Shown here are weighted odds ratios and associated 95% confidence intervals for meeting a sexual partner ≤2km from the original gas leak site by each one-point increase in each sexual stigma type and source (exposure). All participants are included in the main analysis (blue) while only participants living outside 2km are included in the sensitivity analysis (orange), to ensure that results are not just due to the characteristics of participants both living and meeting near the leak site. Adjusted (age, sexual orientation, marital status, education, personal income, and HIV status) main and sensitivity analyses are shown in light blue and light orange, respectively*.

## Discussion

In this cross-sectional study of MSM in Bhopal, men who met male sexual partners near the original leak site of the 1984 gas disaster were more likely to be living with HIV, to be viremic, to have many casual partners, and to engage in transactional sex. We also observed that sexual stigma was a key correlate of meeting a partner near the original leak site. Finally, we observed higher HIV prevalence and viremia among men meeting partners at transportation-related venues, like train stations and bus stops, with the most reported spaces being 1-2 km from the leak site. Taken together, our findings have two major implications: that the areas around the original 1984 gas leak site could be targeted for venue-based HIV control interventions in Bhopal; and that downstream consequences of the 1984 Bhopal gas disaster may continue to impact the spatial distributions of sexual behaviors and HIV burden in MSM four decades later.

One mechanism that could link the 1984 gas tragedy to the modern sexual behaviors of MSM in Bhopal is environmental stigma. Geographic areas which experience industrial contamination are often subject to enduring stigma, with magnitudes depending on distance to the contamination site, the level of poverty of the area, and the speed and adequacy of government response (57–60). The combination of environmental stigma, poverty, and neglect can give rise to or amplify “marginal spaces,” which are subject to social exclusion, reduced visibility, and lower normative control, and can become hotspots for homelessness, sex work, and drug use (61). MSM in India face stigma and social consequences for being identified as MSM and may seek “marginal spaces” to avoid detection when meeting partners (36). The observed link between sexual stigma, engagement with sex work, and meeting a partner near the original leak site, may be because more marginalized MSM choose these locations to avoid detection.

Sexual stigma is linked to HIV and associated risk behaviors through a variety of mechanisms (62). First, sexual stigma in the healthcare setting can reduce the likelihood of HIV testing and viral suppression (63,64), leading to increased potential for onward transmission. Furthermore, stigma can reduce engagement with prevention interventions, such as pre-exposure prophylaxis (PrEP) (64,65). Sexual stigma from familial or societal sources, as well as internalized stigma, may contribute to desires to remain hidden and thus might influence sexual risk-taking and one’s choice of sexual partner or the venue in which they meet them (38,40). A similar effect is seen in people with HIV, who also experience stigma related to their HIV status, and they may also seek the same hidden or stigmatized places to meet sexual partners (66), potentially pairing stigmatized MSM without HIV and MSM with HIV together. The combination of increased willingness to engage in risk behaviors, together with increased exposure to partners living with HIV, creates an environment where the risk of HIV acquisition is heightened. Taken together, the links between HIV, risk behaviors, stigma, and marginal spaces provide a potential mechanism for the heightened burden of HIV among MSM meeting near the 1984 gas leak site.

The idea that HIV transmission is associated with mobility and transportation is well documented in the literature. The HIV epidemic has spread along trucking routes in India and elsewhere (67–69) and even modern analyses identify clusters of new infections and sex work venues near major highways (70,71) and public transport nodes (72). Despite elevated HIV prevalence and viremia, major transportation sites were over 1 km from the leak site, outside the region identified with the highest burden of HIV and prevalence of associated risk behaviors. This suggests that the presence of these transportation-related sites alone does not explain the relationship between HIV and distance to the 1984 site.

Venue-based outreach for HIV-related interventions is a common strategy for populations where no sampling frame exists (73,74). Our results suggest that venues near the original leak site can be targeted, potentially beginning with major transportation hubs and with additional locations chosen after consultation with community informants. Interventions such as condoms and event-driven PrEP are sensible choices for venue-based interventions but even testing and linkage to care interventions in these settings could reach underserved MSM who would not be reached through clinic-based interventions (40,75). The literature on environmental stigma suggests that quick decontamination responses alongside strong governmental support after the next “big event” could curtail the development of environmental stigma and its persistent impact (58–60). In addition to decontamination, economic interventions which reduce entrance into sex work could be considered. Given that the next industrial disaster or similar destabilizing event may have consequences for the burden of HIV among vulnerable populations, the HIV prevention community could be proactive in advocating for early institutional intervention.

This study had a few key strengths. To our knowledge, this is among the first studies to integrate HIV testing, identification of sexual partner meeting locations, and measurement of sexual stigma in a community-based sample of MSM. Sexual partner meeting locations are rarely collected and to our knowledge this is the first study to obtain such meeting locations alongside systematic HIV testing in India. Geospatial studies of HIV burden and risk have predominantly concentrated on the residential location of participants, which may not be appropriate to inform all interventions. Further, our “big events” framework provides the first potential link between an industrial disaster and the spatial distribution of HIV prevalence.

This study also has limitations. We examined distance to the original gas leak site rather than a measure of gas exposure over space, which was not readily available. The gas did not expand equally in all directions, spreading relatively further to the southeast. Distance is therefore an imperfect proxy for gas exposure, though this has also been used in previous studies (19,20). However, because our proposed mechanism linking the 1984 gas disaster and HIV is not direct gas exposure, but rather the creation or exacerbation of marginal spaces facilitated by environmental stigma, it is possible that proximity to the leak site, not just level of gas exposure, could shape perceptions of the surrounding area. Participants could report only up to four areas where they most often met partners, so those classified as not meeting near the site may still have met partners there less frequently. Participants reporting more locations were more likely to report a meeting near the original gas leak site and were more sexually active, and thus the link between meeting near the leak site and HIV could have been due to those associations. However, the spatial gradient of HIV prevalence remained when restricting to participants reporting four locations. Our study was cross-sectional, and thus we cannot assume the direction of causation from our data alone. This has implications for individual-level outcomes, like whether meeting a partner near the leak site was preceded or followed by an HIV infection. At the ecological level, the Union Carbide pesticide plant was placed near a slum and perhaps that area was already stigmatized and a meeting center for MSM engaged in sex work before the gas leak. However, other slums exist in Bhopal, many of which are outside the 2 km radius of the gas leak, so this likely would not fully explain our results. The leak site also lies near Bhopal’s main railway and bus stations, and we cannot fully distinguish proximity to the leak site from proximity to these transport hubs. However, HIV prevalence was highest within 1 km of the leak site, closer than the major transportation hubs, and was elevated near the leak site across venue types. Future analyses could examine similar marginal spaces created or exacerbated by industrial and natural disasters to see if similar links to HIV and stigma are identified.

In conclusion, we identified an area in Bhopal, India where modern MSM meeting male sexual partners faced greater stigma and had a higher prevalence of HIV and associated sexual risk behaviors. We hypothesize that this spatial distribution is linked to the 1984 Gas Disaster, echoing a historical pattern of “big events” exacerbating infectious disease risk in marginalized populations. More disasters and “big events” will occur. If left unaddressed, these events could exacerbate the risk of HIV and other infectious diseases in vulnerable populations for decades to come.

## Data Availability

Data are not publicly available as sharing of sexual partner meeting locations represents risk to this stigmatized population.

