## Supplemental Materials for "Stigma, Place, and HIV: The Legacy of the 1984 Bhopal Gas Disaster Among Indian Men Who Have Sex with Men"

**Supplemental Table 1: Characteristics of men who have sex with men in Bhopal recruited using respondent-driven sampling in 2023-2024, by HIV status (unweighted)**

| **Characteristic**  Median/interquartile range; n/% | **Overall**  **(N=750)** | **With HIV**  **(N=116)** | **Without HIV**  **(N=634)** |
| --- | --- | --- | --- |
| **Socio-Demographics** |  |  |  |
| Age | 25 (21, 32) | 32 (25, 40) | 24.0 (21, 30) |
| Sexual Orientation |  |  |  |
| *Heterosexual* | 216 (28.8) | 22 (19.0) | 194 (30.6) |
| *Homosexual* | 488 (65.1) | 87 (75.0) | 401 (63.2) |
| *Bisexual* | 46 (6.1) | 7 (6.0) | 39 (6.2) |
| Lived in Bhopal >=1 year | 749 (99.9) | 116 (100.0) | 633 (99.8) |
| Marital Status |  |  |  |
| *Never Married* | 531 (70.8) | 64 (55.2) | 467 (73.7) |
| *Currently Married* | 214 (28.5) | 50 (43.1) | 164 (25.9) |
| *Widowed or Divorced* | 5 (0.7) | 2 (1.7) | 3 (0.5) |
| Highest Education |  |  |  |
| *Primary* | 8 (1.1) | 4 (3.4) | 4 (0.6) |
| *Secondary or High School* | 204 (27.2) | 49 (42.2) | 155 (24.4) |
| *Post-Secondary* | 538 (71.7) | 63 (54.3) | 475 (74.9) |
| Earning Pattern |  |  |  |
| *Monthly or Weekly Wages* | 348 (46.4) | 53 (45.7) | 295 (46.5) |
| *Daily Wages* | 84 (11.2) | 30 (25.9) | 54 (8.5) |
| *Intermittently Employed* | 3 (0.4) | 2 (1.7) | 1 (0.2) |
| *Not Employed* | 23 (3.1) | 7 (6.0) | 16 (2.5) |
| *Student* | 292 (38.9) | 24 (20.7) | 268 (42.3) |
| Personal Income |  |  |  |
| *Median (Q1, Q3)* | 40000 (35000, 50000) | 40000 (30000, 45000) | 40000 (35000, 50000) |
| Housing Status |  |  |  |
| *Own* | 557 (74.3) | 96 (82.8) | 461 (72.7) |
| *Rent (Home)* | 114 (15.2) | 16 (13.8) | 98 (15.5) |
| *Rent (Room)* | 69 (9.2) | 3 (2.6) | 66 (10.4) |
| *Other (staying with family, in staff quarters, or in a slum)* | 10 (1.3) | 1 (0.9) | 9 (1.4) |
| Number of Children |  |  |  |
| *0* | 557 (74.3) | 76 (65.5) | 481 (75.9) |
| *1* | 60 (8.0) | 11 (9.5) | 49 (7.7) |
| *2-4* | 133 (17.7) | 29 (25.0) | 104 (16.4) |
| Run Out of Basic Needs (last 12 months) | 126 (16.8) | 24 (20.7) | 102 (16.1) |
| Slept Outside (last 12 months) | 127 (16.9) | 29 (25.0) | 98 (15.5) |
| Access to Smartphone | 740 (98.7) | 114 (98.3) | 626 (98.7) |
| **Sexual Behaviors** |  |  |  |
| Number of Regular Partners |  |  |  |
| *0* | 102 (13.6) | 18 (15.5) | 84 (13.2) |
| *1* | 181 (24.1) | 16 (13.8) | 165 (26.0) |
| *2* | 312 (41.6) | 50 (43.1) | 262 (41.3) |
| *3+* | 155 (20.7) | 32 (27.6) | 123 (19.4) |
| Number of Casual Partners |  |  |  |
| *0* | 8 (1.1) | 1 (0.9) | 7 (1.1) |
| *1* | 44 (5.9) | 1 (0.9) | 43 (6.8) |
| *2-4* | 163 (21.7) | 10 (8.6) | 153 (24.1) |
| *5-9* | 82 (10.9) | 13 (11.2) | 69 (10.9) |
| *10+* | 453 (60.4) | 91 (78.4) | 362 (57.1) |
| Condom Use with Casual Partners |  |  |  |
| *Consistent* | 64 (8.5) | 9 (7.8) | 55 (8.7) |
| *Inconsistent* | 636 (84.8) | 97 (83.6) | 539 (85.0) |
| *Never* | 42 (5.6) | 9 (7.8) | 33 (5.2) |
| *No Casual Partners* | 8 (1.1) | 1 (0.9) | 7 (1.1) |
| Sell Sex | 560 (74.7) | 102 (87.9) | 458 (72.2) |
| Buy Sex | 292 (38.9) | 63 (54.3) | 229 (36.1) |
| **HIV Treatment and Prevention** |  |  |  |
| Ever tested for HIV | 430 (57.3) | 96 (82.8) | 334 (52.7) |
| HIV Test in the Last Year | 98 (13.1) | 10 (8.6) | 88 (13.9) |
| Knowledge of HIV-Positive Status | - | 33 (28.4) | - |
| Currently taking ARVs | - | 32 (27.6) | - |
| Viral Suppression (<200 copies/ml) | - | 78 (67.2) | - |
| **Any Sexual Stigma^** |  |  |  |
| Enacted Healthcare | 148 (19.7) | 32 (27.6) | 116 (18.3) |
| Enacted Family | 124 (16.5) | 32 (27.6) | 92 (14.5) |
| Enacted Society | 122 (16.3) | 22 (19.0) | 100 (15.8) |
| Anticipated Healthcare | 731 (97.5) | 112 (96.6) | 619 (97.6) |
| Anticipated Family | 734 (97.9) | 114 (98.3) | 620 (97.8) |
| Anticipated Society | 734 (97.9) | 111 (95.7) | 623 (98.3) |
| Internalized | 237 (31.6) | 44 (37.9) | 193 (30.4) |
| **Sexual Stigma, Mean (SD)** |  |  |  |
| Enacted Healthcare | 0.15 (0.34) | 0.24 (0.44) | 0.13 (0.31) |
| Enacted Family | 0.16 (0.41) | 0.27 (0.52) | 0.14 (0.39) |
| Enacted Society | 0.15 (0.38) | 0.18 (0.42) | 0.14 (0.37) |
| Anticipated Healthcare | 1.02 (0.41) | 1.00 (0.41) | 1.02 (0.41) |
| Anticipated Family | 1.26 (0.58) | 1.35 (0.60) | 1.24 (0.57) |
| Anticipated Society | 1.09 (0.49) | 1.13 (0.57) | 1.08 (0.47) |
| Internalized | 0.19 (0.37) | 0.24 (0.39) | 0.19 (0.37) |

*^More than “never” (enacted),”very unlikely” (anticipated), and “not at all” (internalized) for any subscale item*

**Supplementary Table 2: Characteristics of Enrolled MSM in Bhopal Stratified by Closest Meeting to Distance to Original Gas Leak Site (Weighted)**

| **Characteristic**  Median/interquartile range; n/% | **<1km**  **(N=47)** | **1-2km**  **(N=216)** | **>2km**  **(N=487)** |
| --- | --- | --- | --- |
| **Socio-Demographics** |  |  |  |
| Age | 25 (24, 35) | 24 (22, 30) | 24.00 (21, 30) |
| Sexual Orientation |  |  |  |
| *Heterosexual* | 8 (35.2) | 42 (30.0) | 166 (48.4) |
| *Homosexual* | 36 (58.7) | 167 (65.5) | 285 (46.2) |
| *Bisexual* | 3 (6.1) | 7 (4.6) | 36 (5.4) |
| Lived in Bhopal >=1 year | 47 (100.0) | 216 (100.0) | 486 (99.7) |
| Marital Status |  |  |  |
| *Never Married* | 30 (67.2) | 156 (75.8) | 345 (73.1) |
| *Currently Married* | 15 (29.6) | 58 (23.6) | 141 (26.7) |
| *Widowed or Divorced* | 2 (3.2) | 2 (0.6) | 1 (0.1) |
| Highest Education |  |  |  |
| *Primary* | 1 (1.8) | 4 (1.0) | 3 (0.3) |
| *Secondary or High School* | 22 (38.6) | 71 (26.8) | 111 (18.6) |
| *Post-Secondary* | 24 (59.6) | 141 (72.2) | 373 (81.2) |
| Earning Pattern |  |  |  |
| *Monthly or Weekly Wages* | 21 (38.7) | 103 (45.7) | 224 (39.7) |
| *Daily Wages* | 10 (17.7) | 34 (12.1) | 40 (6.3) |
| *Intermittently Employed* | 0 (0.0) | 3 (0.9) | 0 (0.0) |
| *Not Employed* | 1 (1.7) | 7 (2.4) | 15 (2.0) |
| *Student* | 15 (41.9) | 69 (38.8) | 208 (52.0) |
| Personal Income |  |  |  |
| *Median (Q1, Q3)* | 40000 (35000, 45000) | 40000 (35000, 50000) | 40000 (35000, 50000) |
| Housing Status |  |  |  |
| *Own* | 42 (87.1) | 171 (76.1) | 344 (66.5) |
| *Rent (Home)* | 3 (6.4) | 28 (12.8) | 83 (15.1) |
| *Rent (Room)* | 2 (6.5) | 14 (9.1) | 53 (15.7) |
| *Other (staying with family, in staff quarters, or in a slum)* | 0 (0.0) | 3 (2.0) | 7 (2.7) |
| Number of Children |  |  |  |
| *0* | 32 (71.1) | 164 (78.3) | 361 (76.2) |
| *1* | 4 (11.6) | 19 (9.2) | 37 (7.8) |
| *2-4* | 11 (17.3) | 33 (12.4) | 89 (16.0) |
| Run Out of Basic Needs (last 12 months) | 9 (12.4) | 56 (18.9) | 61 (10.1) |
| Slept Outside (last 12 months) | 11 (15.0) | 67 (19.6) | 49 (4.5) |
| Access to Smartphone | 45 (96.9) | 213 (98.4) | 482 (98.7) |
| **Sexual Behaviors** |  |  |  |
| Number of Regular Partners |  |  |  |
| *0* | 11 (14.6) | 54 (16.4) | 37 (3.0) |
| *1* | 7 (33.5) | 43 (28.3) | 131 (40.7) |
| *2* | 14 (27.7) | 84 (41.5) | 214 (41.7) |
| *3+* | 15 (24.2) | 35 (13.8) | 105 (14.6) |
| Number of Casual Partners |  |  |  |
| *0* | 1 (2.7) | 0 (0.0) | 7 (3.0) |
| *1* | 0 (0.0) | 6 (8.0) | 38 (16.7) |
| *2-4* | 5 (27.8) | 34 (25.1) | 124 (35.3) |
| *5-9* | 4 (11.6) | 18 (9.9) | 60 (11.6) |
| *10+* | 37 (57.9) | 158 (57.0) | 258 (33.5) |
| Condom Use with Casual Partners |  |  |  |
| *Consistent* | 1 (1.4) | 26 (12.7) | 37 (6.4) |
| *Inconsistent* | 43 (93.9) | 176 (83.8) | 417 (88.5) |
| *Never* | 2 (2.0) | 14 (3.5) | 26 (2.2) |
| *No Casual Partners* | 1 (2.7) | 0 (0.0) | 7 (3.0) |
| **HIV Treatment and Prevention** |  |  |  |
| Ever tested for HIV | 30 (51.7) | 134 (55.4) | 266 (46.4) |
| HIV Test in the Last Year | 8 (11.1) | 42 (13.0) | 48 (6.0) |
| HIV Status: Positive | 15 (21.7) | 40 (14.4) | 61 (9.7) |
| Knowledge of HIV-Positive Status* | 3 (14.5) | 12 (24.2) | 18 (21.6) |
| Currently taking ARVs* | 3 (14.5) | 11 (22.2) | 18 (21.6) |
| Viral Suppression (<200 copies/ml)* | 10 (65.9) | 24 (52.1) | 44 (66.9) |
| **Experiencing Any Sexual Stigma^** |  |  |  |
| Enacted Healthcare | 12 (15.6) | 65 (18.7) | 71 (7.1) |
| Enacted Family | 13 (17.1) | 64 (17.8) | 47 (4.0) |
| Enacted Society | 11 (13.9) | 59 (15.8) | 52 (4.5) |
| Anticipated Healthcare | 46 (98.6) | 207 (98.1) | 478 (99.1) |
| Anticipated Family | 47 (100.0) | 209 (98.7) | 478 (98.9) |
| Anticipated Society | 47 (100.0) | 208 (98.1) | 479 (99.0) |
| Internalized | 20 (29.8) | 94 (32.3) | 123 (14.4) |
| **Sexual Stigma, Mean (SD)** |  |  |  |
| Enacted Healthcare | 0.12 (0.31) | 0.15 (0.35) | 0.04 (0.19) |
| Enacted Family | 0.16 (0.39) | 0.17 (0.41) | 0.04 (0.22) |
| Enacted Society | 0.12 (0.32) | 0.14 (0.36) | 0.04 (0.19) |
| Anticipated Healthcare | 1.01 (0.23) | 1.08 (0.52) | 1.00 (0.22) |
| Anticipated Family | 1.28 (0.48) | 1.28 (0.64) | 1.15 (0.44) |
| Anticipated Society | 1.03 (0.30) | 1.11 (0.57) | 1.05 (0.31) |
| Internalized | 0.17 (0.35) | 0.21 (0.40) | 0.06 (0.20) |

**Among HIV positive respondents*

*^More than “never” (enacted),”very unlikely” (anticipated), and “not at all” (internalized) for any subscale item*

**Supplementary Table 3: Characteristics of Enrolled MSM in Bhopal Stratified by Closest Meeting to Distance to Original Gas Leak Site (Unweighted)**

| **Characteristic**  Median/interquartile range; n/% | **<1km**  **(N=47)** | **1-2km**  **(N=216)** | **>2km**  **(N=487)** |
| --- | --- | --- | --- |
| **Socio-Demographics** |  |  |  |
| Age | 25 (24, 36) | 26 (22, 32) | 24 (21, 32) |
| Sexual Orientation |  |  |  |
| *Heterosexual* | 8 (17.0) | 42 (19.4) | 166 (34.1) |
| *Homosexual* | 36 (76.6) | 167 (77.3) | 285 (58.5) |
| *Bisexual* | 3 (6.4) | 7 (3.2) | 36 (7.4) |
| Lived in Bhopal >=1 year | 47 (100.0) | 216 (100.0) | 486 (99.8) |
| Marital Status |  |  |  |
| *Never Married* | 30 (63.8) | 156 (72.2) | 345 (70.8) |
| *Currently Married* | 15 (31.9) | 58 (26.9) | 141 (29.0) |
| *Widowed or Divorced* | 2 (4.3) | 2 (0.9) | 1 (0.2) |
| Highest Education |  |  |  |
| *Primary* | 1 (2.1) | 4 (1.9) | 3 (0.6) |
| *Secondary or High School* | 22 (46.8) | 71 (32.9) | 111 (22.8) |
| *Post-Secondary* | 24 (51.1) | 141 (65.3) | 373 (76.6) |
| Earning Pattern |  |  |  |
| *Monthly or Weekly Wages* | 21 (44.7) | 103 (47.7) | 224 (46.0) |
| *Daily Wages* | 10 (21.3) | 34 (15.7) | 40 (8.2) |
| *Intermittently Employed* | 0 (0.0) | 3 (1.4) | 0 (0.0) |
| *Not Employed* | 1 (2.1) | 7 (3.2) | 15 (3.1) |
| *Student* | 15 (31.9) | 69 (31.9) | 208 (42.7) |
| Personal Income |  |  |  |
| *Median (Q1, Q3)* | 40000 (35000, 45000) | 40000 (35000, 50000) | 40000 (35000, 50000) |
| Housing Status |  |  |  |
| *Own* | 42 (89.4) | 171 (79.2) | 344 (70.6) |
| *Rent (Home)* | 3 (6.4) | 28 (13.0) | 83 (17.0) |
| *Rent (Room)* | 2 (4.3) | 14 (6.5) | 53 (10.9) |
| *Other (staying with family, in staff quarters, or in a slum)* | 0 (0.0) | 3 (1.4) | 7 (1.4) |
| Number of Children |  |  |  |
| *0* | 32 (68.1) | 164 (75.9) | 361 (74.1) |
| *1* | 4 (8.5) | 19 (8.8) | 37 (7.6) |
| *2-4* | 11 (23.4) | 33 (15.3) | 89 (18.3) |
| Run Out of Basic Needs (last 12 months) | 9 (19.1) | 56 (25.9) | 61 (12.5) |
| Slept Outside (last 12 months) | 11 (23.4) | 67 (31.0) | 49 (10.1) |
| Access to Smartphone | 45 (95.7) | 213 (98.6) | 482 (99.0) |
| **Sexual Behaviors** |  |  |  |
| Number of Regular Partners |  |  |  |
| *0* | 11 (23.4) | 54 (25.0) | 37 (7.6) |
| *1* | 7 (14.9) | 43 (19.9) | 131 (26.9) |
| *2* | 14 (29.8) | 84 (38.9) | 214 (43.9) |
| *3+* | 15 (31.9) | 35 (16.2) | 105 (21.6) |
| Number of Casual Partners |  |  |  |
| *0* | 1 (2.1) | 0 (0.0) | 7 (1.4) |
| *1* | 0 (0.0) | 6 (2.8) | 38 (7.8) |
| *2-4* | 5 (10.6) | 34 (15.7) | 124 (25.5) |
| *5-9* | 4 (8.5) | 18 (8.3) | 60 (12.3) |
| *10+* | 37 (78.7) | 158 (73.1) | 258 (53.0) |
| Condom Use with Casual Partners |  |  |  |
| *Consistent* | 1 (2.1) | 26 (12.0) | 37 (7.6) |
| *Inconsistent* | 43 (91.5) | 176 (81.5) | 417 (85.6) |
| *Never* | 2 (4.3) | 14 (6.5) | 26 (5.3) |
| *No Casual Partners* | 1 (2.1) | 0 (0.0) | 7 (1.4) |
| **HIV Treatment and Prevention** |  |  |  |
| Ever tested for HIV | 30 (63.8) | 134 (62.0) | 266 (54.6) |
| HIV Test in the Last Year | 8 (17.0) | 42 (19.4) | 48 (9.9) |
| HIV Status: Positive | 15 (31.9) | 40 (18.5) | 61 (12.5) |
| Knowledge of HIV-Positive Status* | 3 (20.0) | 12 (30.0) | 18 (29.5) |
| Currently taking ARVs* | 3 (20.0) | 11 (27.5) | 18 (29.5) |
| Viral Suppression (<200 copies/ml)* | 10 (66.7) | 24 (60.0) | 44 (72.1) |
| **Experiencing Any Sexual Stigma^** |  |  |  |
| Enacted Healthcare | 12 (25.5) | 65 (30.1) | 71 (14.6) |
| Enacted Family | 13 (27.7) | 64 (29.6) | 47 (9.7) |
| Enacted Society | 11 (23.4) | 59 (27.3) | 52 (10.7) |
| Anticipated Healthcare | 46 (97.9) | 207 (95.8) | 478 (98.2) |
| Anticipated Family | 47 (100.0) | 209 (96.8) | 478 (98.2) |
| Anticipated Society | 47 (100.0) | 208 (96.3) | 479 (98.4) |
| Internalized | 20 (42.6) | 94 (43.5) | 123 (25.3) |
| **Sexual Stigma, Mean (SD)** |  |  |  |
| Enacted Healthcare | 0.22 (0.41) | 0.24 (0.41) | 0.10 (0.28) |
| Enacted Family | 0.27 (0.50) | 0.29 (0.52) | 0.10 (0.33) |
| Enacted Society | 0.21 (0.43) | 0.26 (0.50) | 0.09 (0.29) |
| Anticipated Healthcare | 1.03 (0.33) | 1.05 (0.56) | 1.01 (0.33) |
| Anticipated Family | 1.36 (0.51) | 1.31 (0.69) | 1.23 (0.53) |
| Anticipated Society | 1.07 (0.40) | 1.11 (0.64) | 1.08 (0.41) |
| Internalized | 0.28 (0.44) | 0.32 (0.48) | 0.13 (0.29) |

**Among HIV positive respondents*

*^More than “never” (enacted),”very unlikely” (anticipated), and “not at all” (internalized) for any subscale item*
